# A minimal and adaptive prediction strategy for critical resource planning in a pandemic

**DOI:** 10.1101/2020.04.08.20057414

**Authors:** Meher K. Prakash, Shaurya Kaushal, Soumyadeep Bhattacharya, Akshay Chandran, Aloke Kumar, Santosh Ansumali

## Abstract

Current epidemiological models can in principle model the temporal evolution of a pandemic. However, any such model will rely on parameters that are unknown, which in practice are estimated using stochastic and poorly measured quantities. As a result, an early prediction of the long-term evolution of a pandemic will quickly lose relevance, while a late model will be too late to be useful for disaster management. Unless a model is designed to be adaptive, it is bound either to lose relevance over time, or lose trust and thus not have a second-chance for retraining. We propose a strategy for estimating the number of infections and the number of deaths, that does away with time-series modeling, and instead makes use of a “phase portrait approach.” We demonstrate that, with this approach, there is a universality to the evolution of the disease across countries, that can then be used to make reliable predictions. These same models can also be used to plan the requirements for critical resources during the pandemic. The approach is designed for simplicity of interpretation, and adaptivity over time. Using our model, we predict the number of infections and deaths in Italy and New York State, based on an adaptive algorithm which uses early available data, and show that our predictions closely match the actual outcomes. We also carry out a similar exercise for India, where in addition to projecting the number of infections and deaths, we also project the expected range of critical resource requirements for hospitalizations in a location.

## 1 Introduction

The World Health Organization (WHO) has declared COVID-19, a disease caused by the novel coronavirus (SARS-CoV-2), as a pandemic. COVID-19 is causing infections and deaths globally on a scale that has not been seen in this century. The virus made a zoonotic transition into humans and then continued person to person transfer. ^1^ Analysis of the 1918 Spanish flu suggests a need for timely action from the Governments. ^2,3^ With no vaccines or treatment options, prevention via social distancing and city or nation-wide lockdowns have become the catchword for containing the spread of infections. These restrictions on the movement of people of course have adverse effects on the economies. There have been several efforts to model the spread of the pandemic ^4–6^ to understand the gravity of the situation that one is facing ^7^ as well as to suggest travel restrictions ^8,9^ and to guide policy measures ^10–12^ on the extent of testing or the implementation of the lockdowns.

Further, the unexpected surge in patients requiring hospitalizations and intensive care is leading to the collapse of health care systems globally. Gearing up for providing the health care supplies including critical equipment such as ventilators requires an accurate estimate of the number of infections. But the key information that is required for this modelling, which is the number of infected people at any time, is in itself marred by uncertainties. There has been an acute shortage of testing kits required for extensive screening. As a result, although symptomatic patients may have been traced or tested, a significant fraction of the global infections are believed to have been through asymptomatic patients. ^13^ Although a pandemic such as COVID-19 appears as a once-in-a-century event, the 21st century has already seen a few others which drew very close: SARS in 2003, swine flu in 2009, MERS in 2015. Thus, having tools to quickly plan for the critical resource requirements is important.

The nature of transmissible diseases such as COVID-19 is that the number of infected people grows exponentially. The leaders of many countries have made a sudden switch from complacency to active mitigation policies, as if they were caught by surprise while expecting linear trends. The nature of the numerics associated with the long-term pandemic trajectory evolution is that the errors in estimation also increase exponentially, no matter how detailed the model is. An early prediction of a pandemic that forecasts the evolution for many months will accrue errors very quickly, and a model that is developed much later in the evolution will only be relevant for *post facto* analyses, and not for policy decisions or disaster management.

Therefore, the models should be designed to be adaptive, such that their projections are constantly corrected. They should also be simple enough to be interpretable by the various stakeholders (with differing abilities to understand the detailed models) involved in the management of the pandemic. Further, due to the differences in the allowed freedom of movement or the rigour of policy implementation in various states or provinces in different countries, the predictions should be recalculated.

The focus of this work is to set up a framework that can be useful to estimate the need for critical resources such as hospital beds and ventilators, the need for which changes with time and with the region (province, state or district) of interest. *Inter alia*, we also make predictions based on early data for Italy and New York state, update them, and demonstrate good agreement between the subsequent evolution of the pandemic and our predictions. Our approach can be summarized as follows: The COVID-19 data from most countries suggests that, especially in the growing phase of the pandemic, the number of active cases and the number of hospitalizations are both proportional to the total number of infections: approximately around 70-90 % and 20-30%, respectively. This conclusion is arrived at by eliminating “time” as a variable, and focusing only on the above-cited ratios. Moreover, it is quite easy to update our estimates on a weekly basis.

## 2 Results

### 2.1 Universal trends in infection and death data

A simple law that can capture the spread of infections is

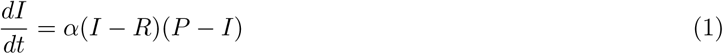

where *α* is the rate of transmission, *I* is the number of infected people, *R* is the number of people who recover from infection and *P* is the total population. Further details can be added by noting that *I* is the sum of symptomatic and asymptomatic patients, and that *α* differs for the two groups and across communities depending on their social contact structure or policies of various degrees of isolation. However, this is not done here.

When the infections are in a growing phase, the data suggests that *I* ≫ *R*, which in turn suggests that *I* can be used as a proxy for the number of *active* infections. Coming to the focus of this work, we sugguse that the number of active infections can be the basis for critical resource planning during the evolution of the pandemic. Also, regardless of the scale of the pandemic, it remains manageable only when *P* ≫ *I*. This allows one to approximate Eq. 1 as

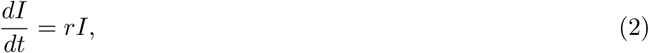

where *r* is the effective rate of the spread of infection. The parameter *r* depends on the infection under consideration. During the early days of the pandemic, when detailed biological understanding of the disease spread is still elusive, it can be a challenge to estimate the parameter *r* accurately. One approach is to estimate *r* using the time-series of the disease evolution. However, analyzing the raw time series is problematic, because *I* is an integer-valued variable, and further, its values would be small at the outset. Moreover an analysis of the raw time series data from several countries reveals that *there is no universal value for r* across countries. This is illustrated in Figure:1A. Even the cumulative value of *I* does not reveal any universal trend (see Figure:1B). Therefore it is necessary to adopt a different approach.

**Figure 1:**
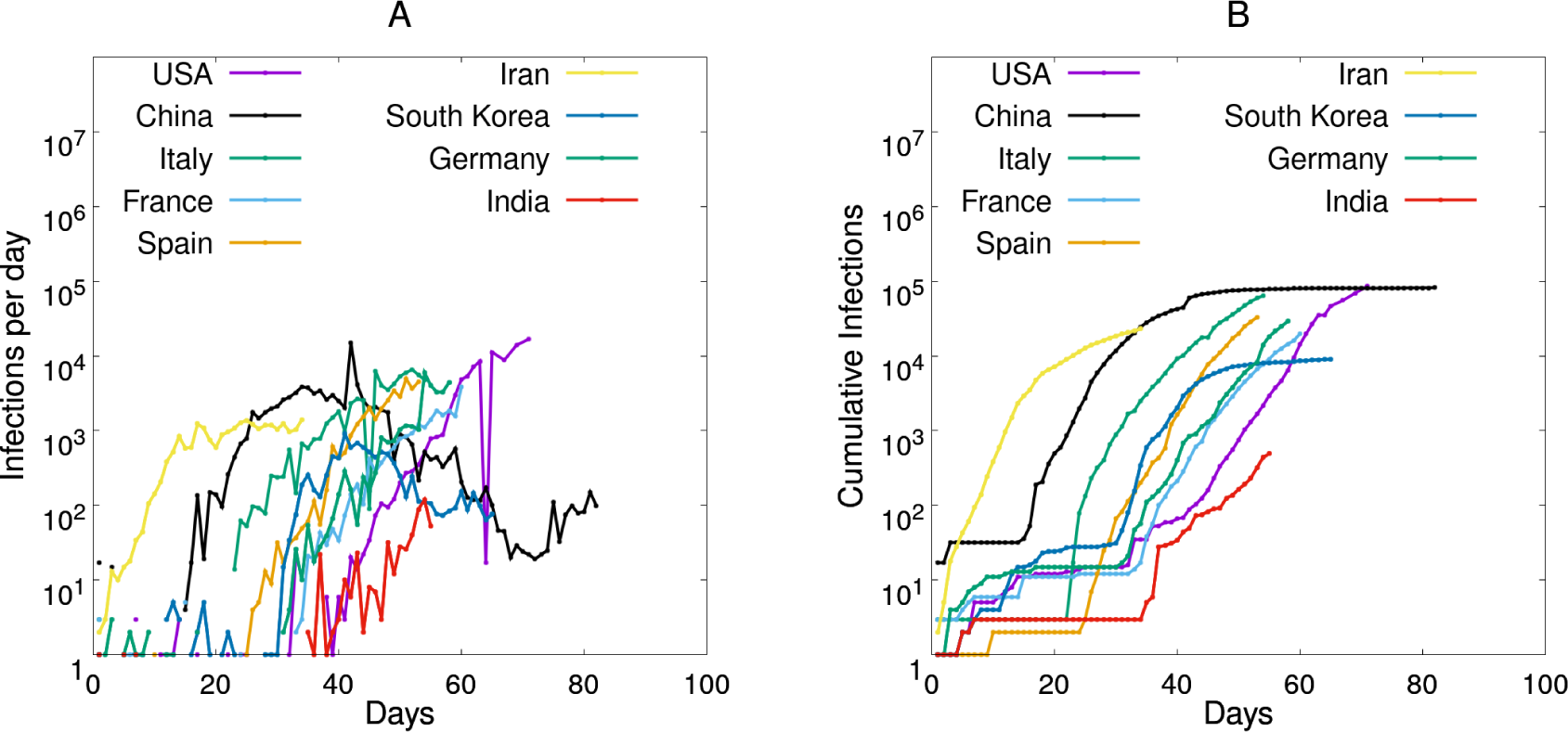
Time series has fluctuations. Country-wise time series data for (A) daily new infections (*dI/dt*)and (B) cumulative infections(*I*). Since the chance of infection and incubation time are stochastic, and data is recorded on a daily basis, *I* fluctuates. These fluctuations may be country-dependent and constrained by other factors such as the frequency of testing, specially in early days of the pandemic. To obtain a better evolution we instead focus on the (*dI/dt*) versus (*I*) which removes systematic errors

Our approach is to *eliminate time as a variable*. By doing so, we found three universal trends, one partly expected and the others not so immediately apparent:

1. The infection rate is proportional to the number of current infections (Figure:2A). In principle this is expected from Eq 2. In practice this finding is interesting because the rates from all countries are comparable, especially during the early days of the spread of infection before any policy changes are implemented. This also underscores the fact that the biology of the disease is similar from one country to the other, without further mutations in the virus or differences in immunity levels of the different populations. This commonality across the data from different countries has been previously highlighted by many.^14^
2. Interestingly, there is also another universal pattern across the countries, where the rate of deaths caused by the infection is proportional to the number of deaths (Figure:2B). Further, the fluctuations in this data are lesser in extent compared to that in Figure:2A, a feature that we will exploit later. It is not immediately apparent why the rate of deaths should be proportional to the cumulative deaths. One possible explanantion is that the number of deaths depends directly on the number of infections.
3. The cumulative number of deaths at any time is directly related to the number of overall infections(Figure:3A). Considering the wide variation across countries in the duration of the hospitalization and critical care, it is not apparent why this should be so. However, the similarity in the data from across the countries is striking, and we utilize it in our model.

**Figure 2:**
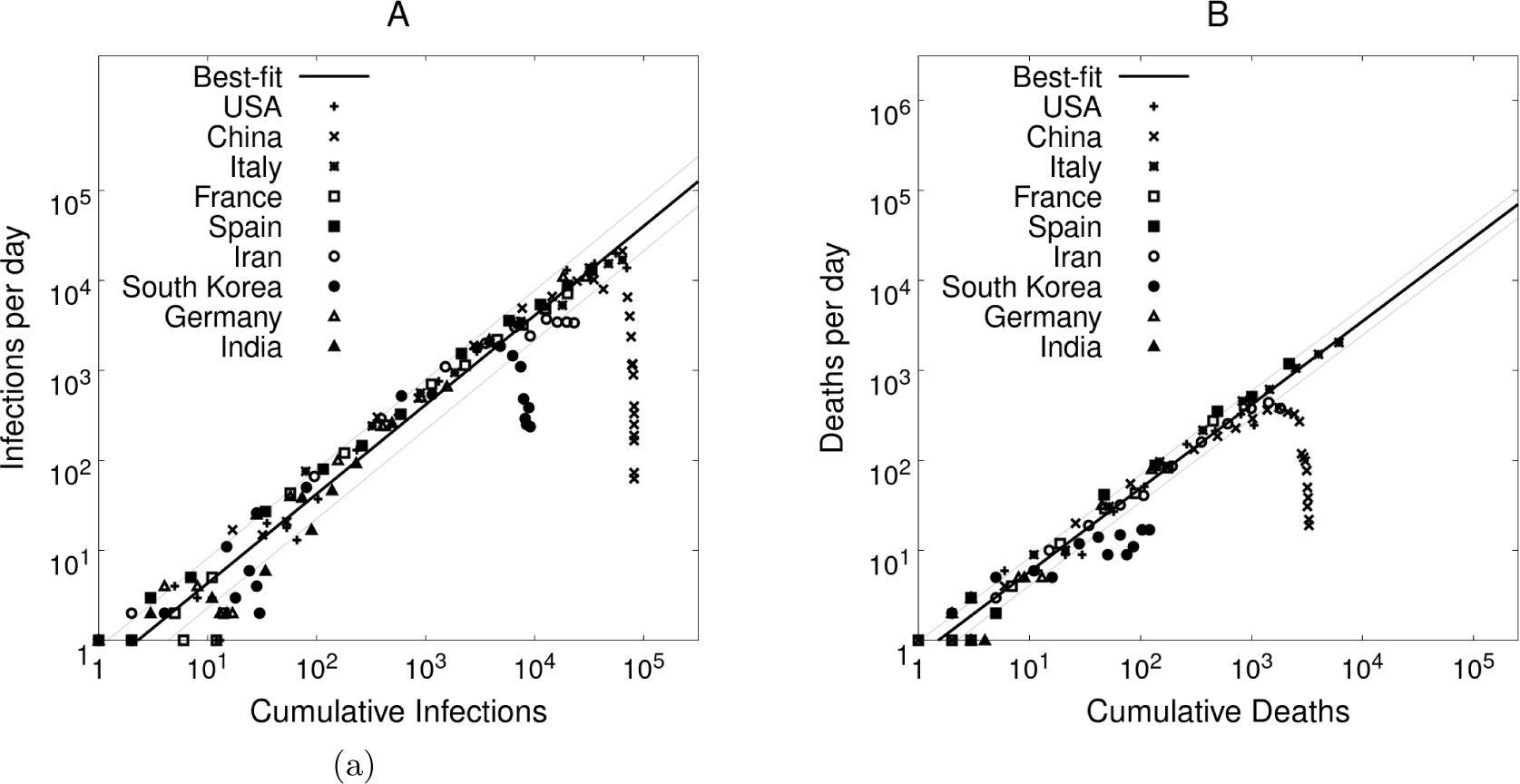
Universal trend in infections and deaths. The rates at which the number of infections (*I*) and number of deaths (*D*) change are explored using the COVID-19 data from many countries. (A) *dI/dt* versus *I* (B) *dD/dt* versus *D* show comparable behaviour across these countries which can be exploited in our predictions. The data from a 3-day average was used since the number of daily new infections fluctuates significantly. The standard deviation from the best fit curve in the infection plot is 0.276 and in the deaths plot is 0.153

**Figure 3:**
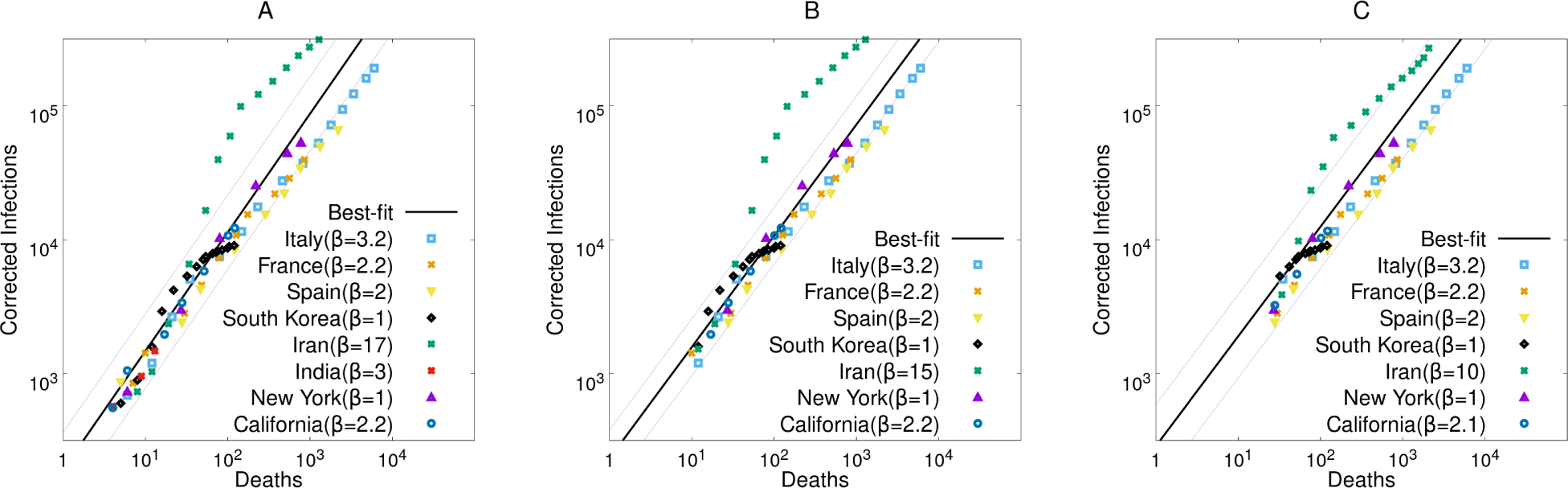
Using the I-D plot for estimating number of infections. These plots depict the universal relation between I and D, barring a corrective factor. This uncertainty factor that was calculated from the COVID-19 infection data of South Korea as a reference can be used for estimating infections when the number of deaths are known. 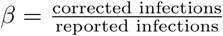 was assumed to be 1 for South Korea. At any time in the evolution of the pandemic shown on this *I* versus *D* graph, using the known number of deaths in that country, the number of infections were rescaled to match with the infections in South Korea and this factor was used as *β*. The figure illustrates the procedure used for *β* when deaths are slightly lower than 10, equal to 10 and 20.

### 2.2 Number of deaths as an additional and surrogate marker for number of infections

As noted earlier, it is not easy to scale up the number of test kits and to arrange tests in large populations. Thus the official count of the number of infected individuals at any given time is far from being very precise. An alternative way of estimating the number of infections is via monitoring of the the number of deaths. The key idea is that the fraction of infected people who may die are most likely to be in hospitals and thus a fraction of the reported infection by the state. While this fraction might depend on country and quality of testing, it is more likely that an infected person who may die due to other health complications will be in hospital. To do this, we incorporate a scaling factor to account for the extensiveness of the testing. It is believed that, to date, the most extensive screening tests have been performed by South Korea, detecting most of the symptomatic and asymptomatic infected individuals. Thus, using the data from South Korea as a reference standard, the deaths versus infections curve has been readjusted as seen in Figure:3A,B, and C. The scaling factor required for each data set is used as an uncertainty factor *β*, which accounts for variations in the criteria for conducting screening tests. Thus, assuming the risk profiles of infected individuals are constant across countries, the number of observed deaths can suggest another way of estimating the number of infected.

We use two different measures of the number of infected: one by the number of individuals who test positive and two by the estimated number of infected individuals by studying the number of deaths. It is understood that the assumption comes with limitations: firstly the testing in South Korea may not have completely captured all the infections but only a majority; secondly the number of deaths per 1000 infections may be different if individuals of a higher median age (for example in Italy) are infected compared to individuals of a slightly lower median age (for example in Germany). However, remembering that the objective is to plan for the resources needed for the individuals who are currently infected, the lower death rate is likely to predict the number of infections on the safer side as far as resource planning is concerned. The estimate of infections from these two different approaches will be used to set the limits on the number of infected individuals, and the resources required.

### 2.3 Piecewise-prediction with adaptive re-training

The preceding subsection addressed the difficulties in measuring *I*, the number of infections. In this subsection we discuss the estimation of *r*, the rate at which the infection spreads. A typical way to estimate *r* is to fit the time series of *I*(*t*) by an exponential. As pointed out earlier, when *I* is small, fitting a time series to an integer-valued (and possibly noisy) variable is problematic. Instead, we use the slope of *dI/dt* versus *I* (Figure:2A) to estimate *r*. Instead of applying Eq 2 exactly with all the attendant uncertainties in *I* and *r*, we use a piecewise strategy. A simple discretization of this in time steps of Δ*t* gives *I*(*t* + Δ*t*) = *I*(*t*) + ⟨*r*⟩_*t,t*+Δ*t*_ · *I*(*t*). It was seen from the COVID-19 infection data from several countries that as the number of daily new infections fluctuates considerably, the trends from the data can be captured only by following the data over a few days. We work with a Δ*t* = 7 days, since inferring *r* from the data drawn over a shorter duration or making a prediction more fine-grained than this do not seem to be relevant from a practical point of view.

The multiplicative factors ⟨*r*⟩_*t,t*+Δ*t*_ estimated week by week, using the universal patterns observed in Figure:2B, are given in Supplementary Table 1. We make two predictions for *I*(*t*): one using the reported infections from the country, and another using the reported deaths. The scaling factor *β* is re-calibrated once a week in an adaptive fashion, using the data until that week. In essence, the exponential function of time has been replaced by a geometric series for each week.

**Table 1:**
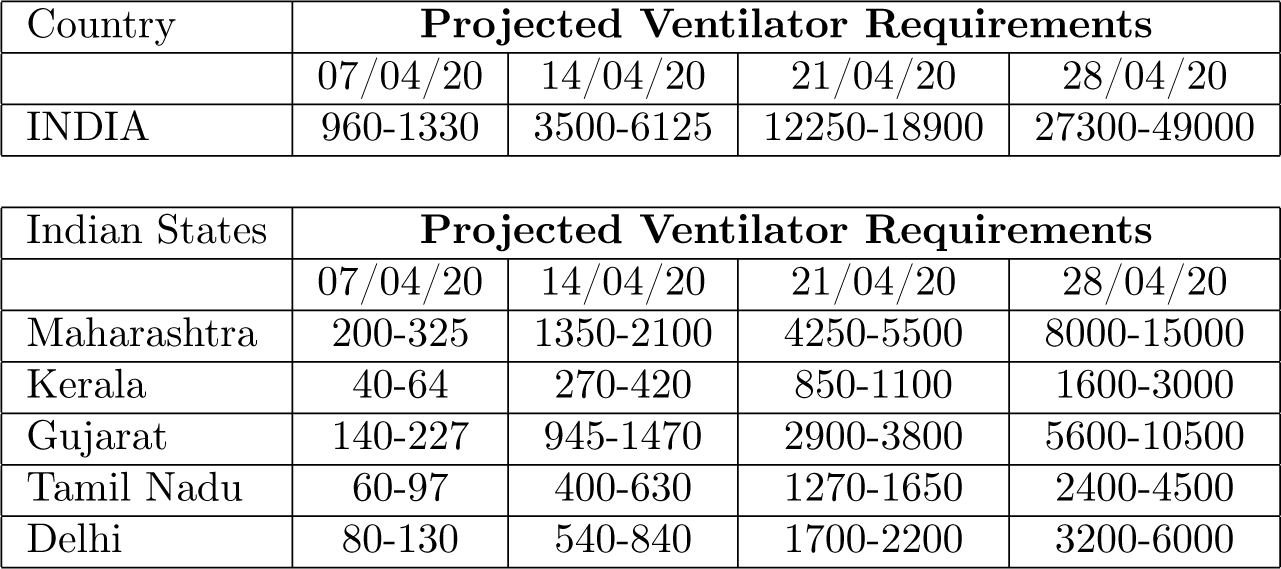
Projected ventilator requirements for India and top 5 worst affected states as of 31st March 2020. The projections have been made for the upcoming 4 weeks using the approximate relationship that the mortality is 20% of patients who are on ventilators

| Country | Projected Ventilator Requirements |  |  |  |
| --- | --- | --- | --- | --- |
|  | 07/04/20 | 14/04/20 | 21/04/20 | 28/04/20 |
| INDIA | 960-1330 | 3500-6125 | 12250-18900 | 27300-49000 |

| Indian States | Projected Ventilator Requirements |  |  |  |
| --- | --- | --- | --- | --- |
|  | 07/04/20 | 14/04/20 | 21/04/20 | 28/04/20 |
| Maharashtra | 200-325 | 1350-2100 | 4250-5500 | 8000-15000 |
| Kerala | 40-64 | 270-420 | 850-1100 | 1600-3000 |
| Gujarat | 140-227 | 945-1470 | 2900-3800 | 5600-10500 |
| Tamil Nadu | 60-97 | 400-630 | 1270-1650 | 2400-4500 |
| Delhi | 80-130 | 540-840 | 1700-2200 | 3200-6000 |

The results from adaptively predicting the number of COVID-19 related deaths from Italy are illustrated in

Figure:4A. Initially, the data of the number of deaths from week 1 was used to predict for weeks 2 to 4. Later, using the data for week 2, predictions were made for weeks 3 to 5. As it can be seen, the prediction at week 4 improved after retraining using the data from week 2. A similar adaptive prediction for the number of COVID-19 infections in Italy is shown in Figure :4B, using death and infection count as two independent estimates. The predictions for death were more accurate than the predictions of infections. This could be theoretically possible if the number of tests increase linearly with time, while the infections grow exponentially, thus increasing the gap of untested individuals. Many countries which initially allowed testing or hospitalizations during the initial phases of the pandemic have made the criterion stricter as the resource gap increased over time. Overall, it is clear that the trends in the number of infections can be captured with this simple approach. The model is simple enough that it can be repeated easily for making the predictions of the COVID-19 cases in any other state or country, each of which can be adapted on a weekly basis. We show these results for the New York state (Figure:5) and India (Figure:6). One can see the same pattern as for the data from Italy that the predictions of COVID-19 deaths in New York improves with the iteration. The prediction of infections continues to be an higher than the reported infections, just as in for the data from Italy. The numbers of deaths and infections predicted for India are still low, and at this point seem to be in accordance with the predictions.

**Figure 4:**
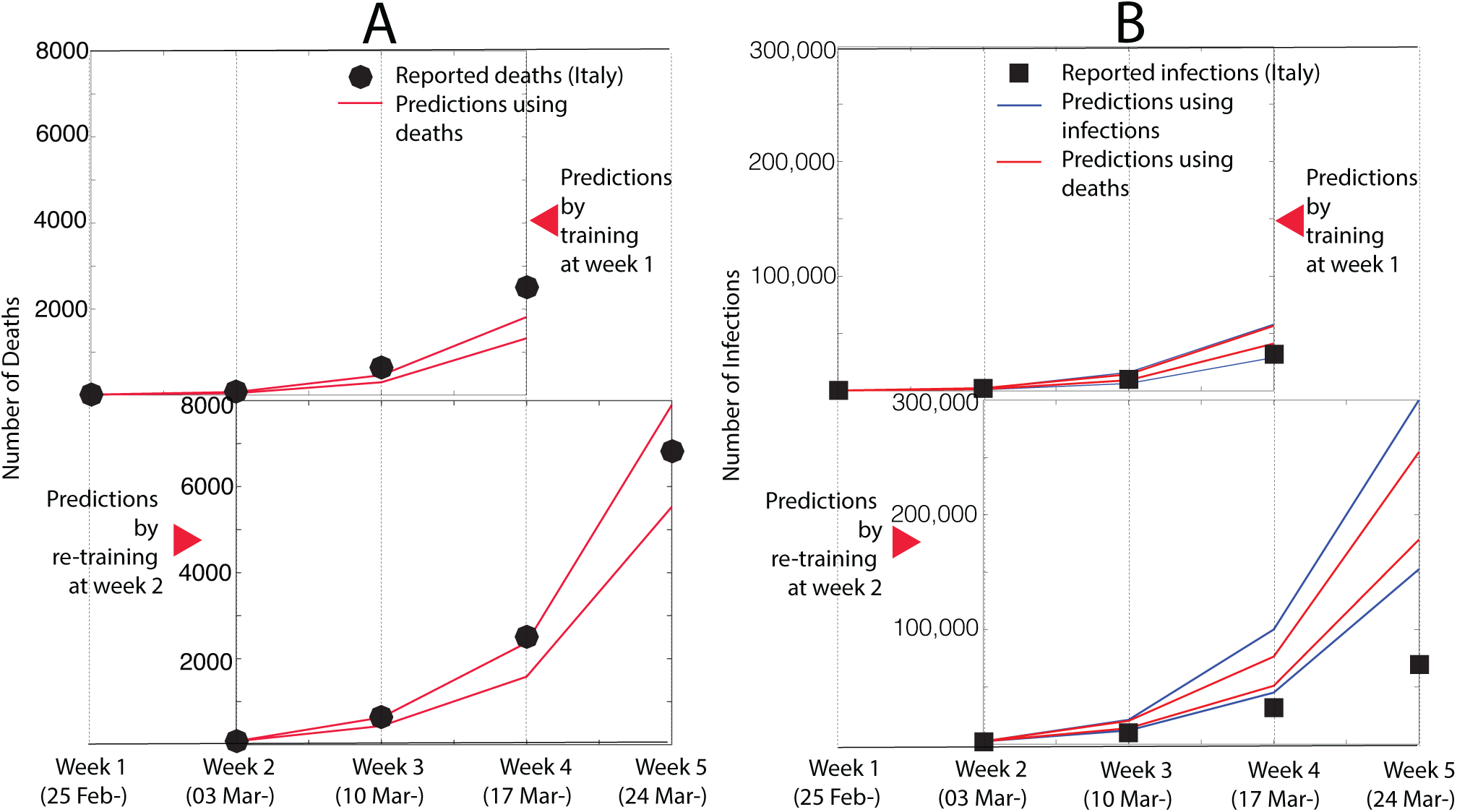
Adaptive predictions for Italy. An Illustration of the adaptive predictions for (A) the number of deaths and (B) the number of infections due to COVID-19 in Italy. In both these cases, the top panel uses the the data from week 1, and predictions for weeks 2 to 4. At the end of week 2, by taking note of the reported data, the predictions for weeks 3 to 5 are updated. The reported number of deaths and infections are also shown. In comparison, the prediction of the number of deaths was more accurate. It should be noted that number of deaths also gives one a better estimation of the critical resources required. This could potentially be because the infections grow exponentially and the testing resources grow linearly thus increasing the gap in the reported infections.

**Figure 5:**
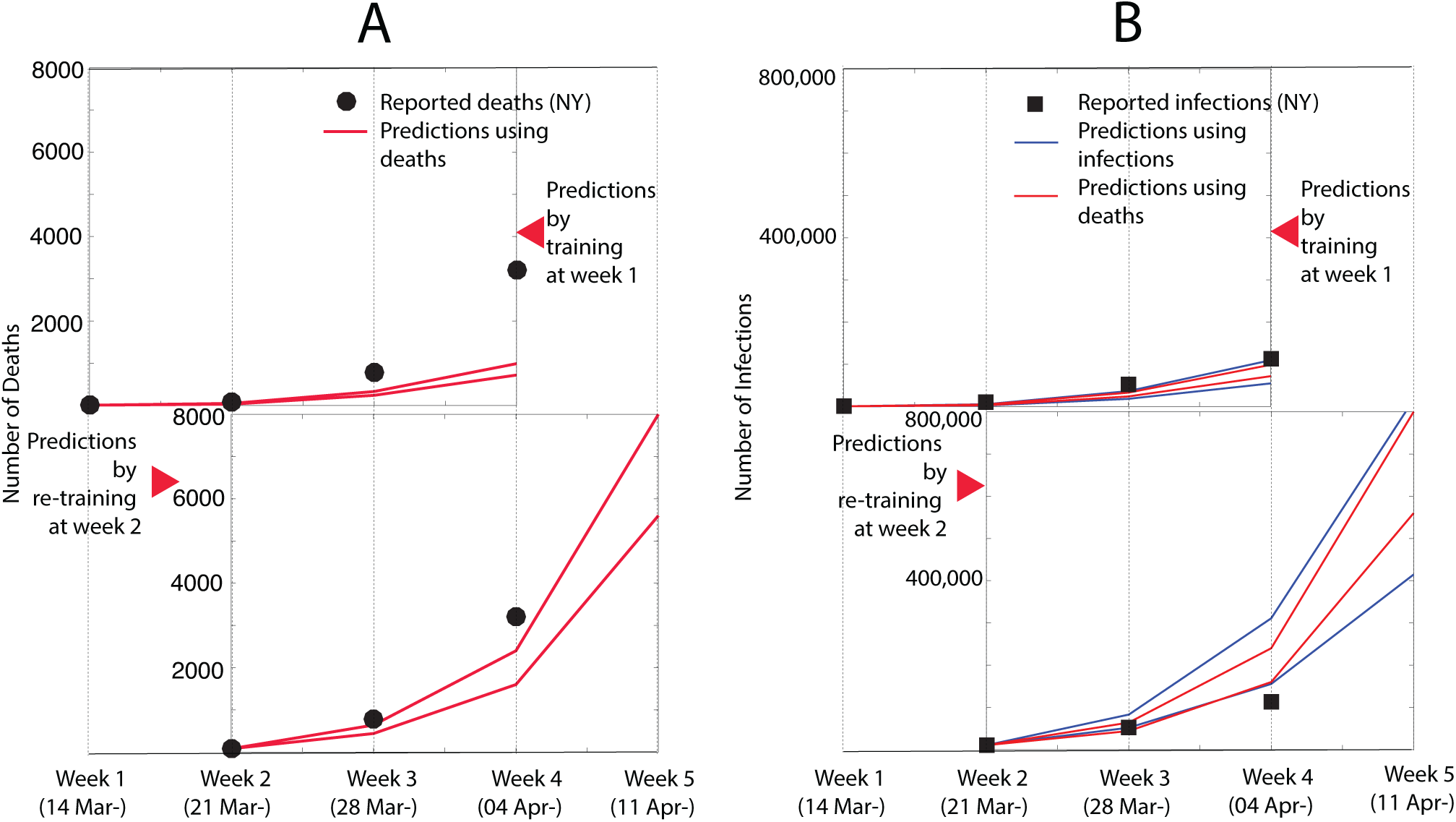
Adaptive predictions for New York state. The adaptive predictions were performed for the data from New York state, just as described in Figure:4.

**Figure 6:**
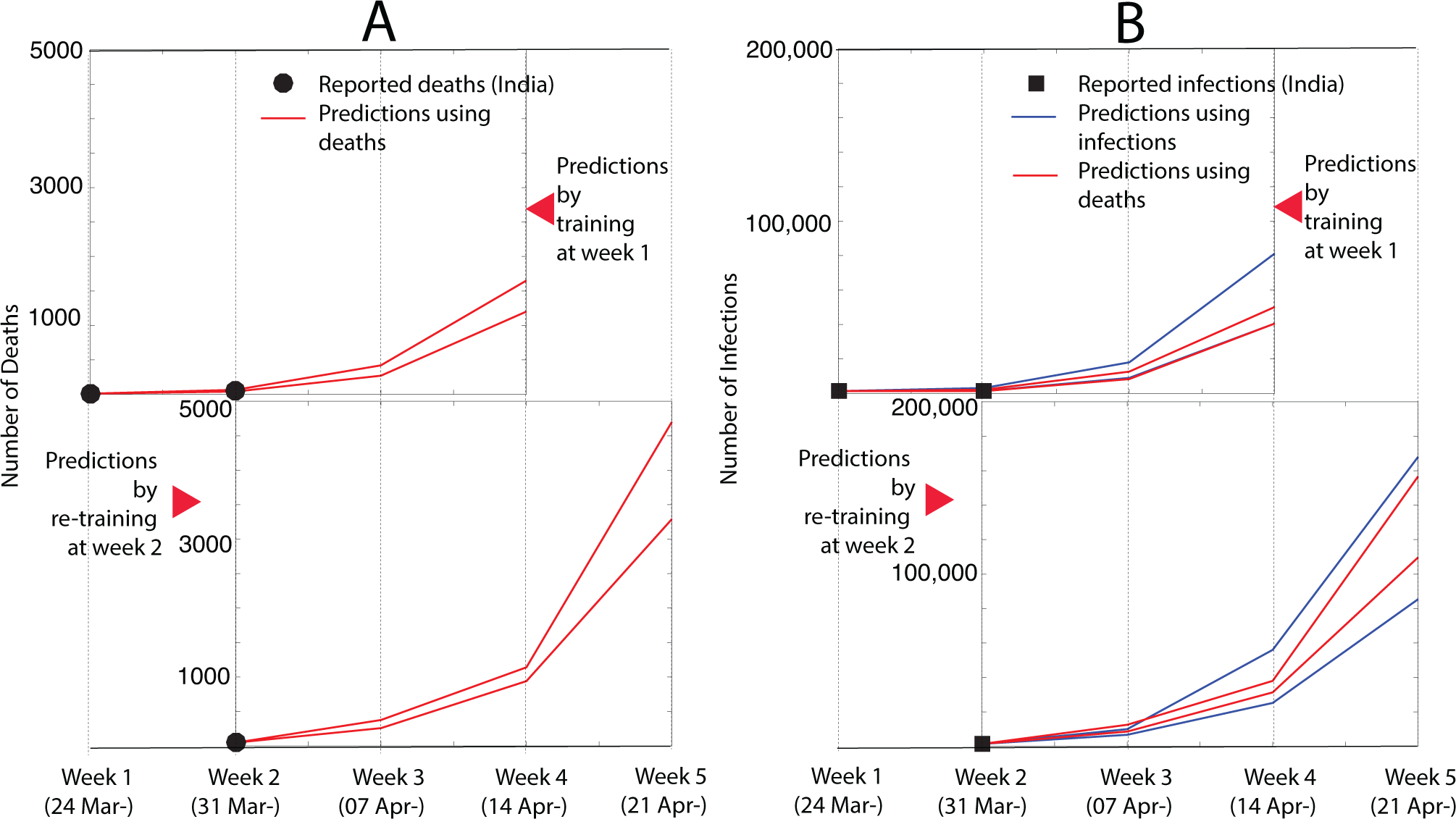
Adaptive predictions for India. The adaptive predictions were performed for the data from India, just as described in Figure:4 and Figure:5

### 2.4 Effects of a lockdown

A lockdown can cause two changes in our model. First, depending on the levels of restrictions on movement such as social distancing, ⟨*r*⟩_*t,t*+Δ*t*_ will reduce or even drop to zero. The ⟨*r*⟩_*t,t*+Δ*t*_ in the weeks following important restrictions on movement from different countries was also used as a lower bound on how fast the new infections can decrease. Second, when a country imposes a lockdown by imposing restrictions on international or inter-state movement, the different physically separated regions will follow their own independent growth trajectory. The weekly growth factor to be applied to each specific state or province will depend upon the number of infected in the province at the time of the decision, with some regions lagging behind the others by weeks.

One thing we learnt from COVID-19 is that each country or province, before it becomes a transmission vector for other places, has a lead of several weeks or months over those places. The lead of course depends on the extent of people travelling between these places. A disease that was once confined to a specific region, diffuses locally and jumps to distant locations almost like a jump-diffusion process. There is a self-replication phenomenon at different scales, wherein the universalities in the patterns between the nations and at a national level, now are reflected between the individual states or provinces. This observation can be used by the governments of these provinces to plan for the worst-case scenario. Each pandemic is different in its ability to persist, infect, and diffuse through the population. While the medical knowledge and critical equipment required may differ from the earlier pandemics, as long as the universalities discussed in this work are seen in the data, one can certainly learn from other countries, states or provinces that experienced the same pandemic a few weeks or months earlier.

### 2.5 Outlook for disaster management

Various governments may wish to use the number of infections as a metric to test the effectiveness of their containment measures, because the fatalities in a pandemic depend not just on the nature of the virus, but on many more variables, such as patient history, quality and availability of healthcare, and preparedness. Thus both the number of infections and deaths need to be predicted and validated. The rapid spread of COVID-19 has resulted in a sudden spurt in the demand for critical health services. The planning, production and distribution of these resources in itself require lead times of several weeks to months. Hence, there is a need to have a disaster management plan that can predict the resource requirement in the weeks to come. But a forecast done based on noisy inputs from the early days of infection may turn out much a like a long-term weather prediction. While epidemiological models often attempt to predict the rate of infection spread, the requirements for critical patient care are often not directly modelled. The predictive model developed here is distinctive in using fatalities (*D*) as the key parameter rather than the number of reported infections (*I*). This key observation, coupled with the use of a phase plot (corrected infections versus deaths) allows us to provide projections for the critical health-care sector. We have performed this exercise for India and for indivdual states (see Table 1). ^15^ Table 1 lists the projected number of ventilators required in each state as well as the gross national estimate for a period of four weeks beginning from April 7th, 2020.

## 3 Conclusions

In this work we aimed to provide tools for critical resource planning in the wake of a pandemic. We do this by introducing three attributes that are not common in epidemiological modeling: We used the number of deaths as an additional and surrogate marker for the otherwise uncertain number of infections. Given the data is stochastic, and with several unknowns, we focused on making weekly predictions based on a geometric series with piecewise adaptive ratio. Adapting the geometric ratio week by week serves two objectives: (i) smoothing out the daily fluctuations in data, and (ii) bridging the gap between predictions made using early data, and current data. The simplicity of the formalism, and its data-aware design permit us to make adaptive predictions for the next few weeks, starting at any time. Although COVID-19 is a rare pandemic, the quick, simple and adaptive principles laid out here, embracing the limitations of testing and drawing on the universalities of the progression, should be relevant for any pandemic spreading through person to person contact.

## Data Availability

All data have been sourced from open-source locations.

https://mesoscalelab.github.io/covid19/

https://www.worldometers.info/coronavirus/

## Acknowledgement

We acknowledge Dr. Rahul Tyagi and Dr. Vinay Kariwala valuable suggestions. We sincerely acknowledge discussions with Prof. G. U. Kulkarni, Prof. K. R. Sreenivas, Dr. Sunil D. Sherlekar and Prof. M. Vidyasagar. We thak JNCASR and IISC for supporting this research via an initiative of the office of the Principal Scientific Advisor, Govt of India.

## Declaration of Interests

The authors declare no competing interests.

## Author Contributions

A.K and S.A designed research; M.K.P, S.K, S.B, A.K, A.C and S.A performed research; M.K.P, S.K, A.K, and S.A analysed data; and M.K.P, A.K, and S.A wrote the paper.

## Notes

### Competing Interest Statement

The authors have declared no competing interest.

### Funding Statement

We can thank Office of Principal Scientific Advisor to GoI and JNCASR for their support.

